# Predicting animal to human translation: A proof of concept study using qualitative comparative analysis

**DOI:** 10.1101/2022.01.31.22270227

**Authors:** Cathalijn H.C. Leenaars, Steven Teerenstra, Franck L.B. Meijboom, André Bleich

## Abstract

Drug development suffers from high attrition rates; promising drug candidates fail in clinical trials. Low animal-to-human translation may impact attrition. We previously summarised published translational success rates, which varied from 0% to 100%. Based on analyses of individual factors, we could not predict translational success.

Several approaches exist to analyse effects of combinations of potential predictors on an outcome. In biomedical research, regression analysis (RGA) is common. However, with RGA it is challenging to analyse multiple interactions and specific configurations (≈ combinations) of variables, which could be highly relevant to translation.

Qualitative comparative analysis (QCA) is an approach based on set theory and Boolean algebra. It was successfully used to identify specific configurations of factors predicting an outcome in other fields. We reanalysed the data from our preceding review with a QCA. This QCA resulted in the following formula for successful translation:

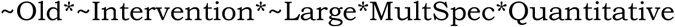

Which means that within the analysed dataset, the combination of relative recency (∼ means not; >1999), analyses at event or study level (not at intervention level), n<75, inclusion of more than one species and quantitative (instead of binary) analyses always resulted in successful translation (>85%). Other combinations of factors showed less consistent or negative results. An RGA on the same data did not identify any of the included variables as significant contributors.

While these data were not collected with the QCA in mind, they illustrate that the approach is viable and relevant for this research field. The QCA seems a highly promising approach to furthering our knowledge on animal-to-human translation and decreasing attrition rates in drug development.

## INTRODUCTION

While the debate on the relevance and acceptability of animal experimentation remains polarized [1-4], animal experiments are still hard to avoid in the process of new drugs reaching the market. However, the predictive value of animal experiments has limits, and poor translation from animal experiments to humans may contribute to the high attrition rates in drug development [5]. Explaining attrition can contribute to more efficient drug development, which is one of the reasons why we analyse translational success. Another one is animal welfare; we cannot defend using animals for translational experiments that do not provide relevant information. While the most common approach to evaluating translation is mechanistic and qualitative, we started focussing on quantitative studies in a scoping review of reviews [6]. In that review, we observed translational success rates from 0% to 100% (median: 64%; interquartile range (IQR): 44-79%). To identify factors contributing to translational success, we visualised these data by several potentially predictive factors, which are explained further below. Relevant for this paper are: definition type (binary vs. continuous definitions of translation), unit of analysis (were the results analysed at the event, intervention or study level), species, the number of included observations (events, interventions or studies), and the year of publication. There was no apparent relationship between any of these individual factors and the percentage of translational success. However, the effects of combinations of these potential factors on translational success could still be relevant. We thus performed additional analyses on our previously-collected data, which are described in this paper.

The following five factors were further analysed based on their theoretical relevance: publication age, unit of analysis, analysis size, inclusion of multiple species and type of definition for translation (binary vs. continuous). Publication age was included as the state of science and the quality of animal models are thought to improve over time, and because animal-to-human translation is getting more attention in the last decade, which may result in improvements.

The unit of analysis was previously extracted as a categorical variable with 3 possible values: event, intervention or study. Events were mainly specific adverse events observed in animals, humans or both, where the observed translation (e.g. the percentage of adverse events observed in both animals and humans) depends on study size and chance; larger studies have a larger chance of picking up rare events [7]. Interventions were mostly specific drugs, where certain groups of interventions may translate better than others [8], and the observed translational success rate depends on the sampling, e.g. which group of drugs was analysed. Analyses at the level of individual studies show translation as results that corresponded between animals and humans, which makes the observed translational success rate depend on multiple factors, including population and experimental design of the compared studies [9].

The number of included observations was counted at the level of the unit of analysis, and could thus reflect a number of events, interventions or studies. It was included in the current analyses as a proxy for power, as underpowered studies can result in erroneous conclusions which may impact translation [10]. While some authors argue that species differences introduce uncertainties that seriously limit their validity [11], investigating multiple species can at least theoretically improve translation, as successful transfer of a first species barrier may be predictive of crossing a second. The definition type for animal to human translation could be binary, i.e., there was successful translation or there was not, or continuous, which could refer to a percentage success, a correlation coefficient between animal and human data, a percentage overlap in confidence intervals, etc. Binary definitions can of course be expressed as percentages success, but the type of definition may impact the observed translational success.

As multiple roads lead to Rome, multiple combinations of these factors may lead to translational success. In scientific terms, there possibly is causal complexity; comprising equifinality (i.e., there are multiple routes to success) [12] and conjunctural causation (i.e., combinations of factors may be involved instead of individual factors [13]). Besides, causation may be asymmetrical [14]; while the presence of a factor may contribute to success, its absence does not necessarily result in failure (and vice versa). Thus, we have a configural research question: “Which factors, individually or in combination, are necessary or sufficient for successful animal-to-human translation?” To analyse effects of multiple potential predictors on an outcome, regression analysis (RGA) is common. However, RGA is not specifically suitable for research questions comprising multiple interactions and specific configurations (≈ combinations) of predictors. Qualitative comparative analysis (QCA) is an approach developed for configural research questions [15]. It is based on set theory and Boolean algebra. QCA is increasingly used to identify specific configurations of factors predicting an outcome in other fields [16, 17]. We reanalysed the data from our preceding review with a crisp-set QCA (csQCA) [18]. To test the added value of this QCA-approach, we compared it with a classical regression analysis (RGA).

## METHODS

### Data collection and selection

We reanalysed the data published in our systematic scoping review [6] for this study. This preceding scoping review was an umbrella review of reviews that addressed animal-to-human translation quantitatively, comparing the results of studies including at least 2 species with one being human. Data were extracted from the included publications to Microsoft Excel. When an included paper described multiple studies or analyses on different data, all those compliant with the inclusion criteria were included as a separate “case” into our analyses. When the original authors did not express translation as a percentage, but provided the data needed to do so, we calculated the percentage and added it to the respective case.

From these already published data, we selected the following factors as theoretically relevant for further combined analyses (as explained in the introduction): definition type (binary vs. continuous), unit of analysis (event, intervention or study), species, the number of included observations and the year of publication. Cases with missing data for any of the analysed factors were excluded from the analyses (numbers are mentioned in the results).

All analyses were performed in R [19], version 4.0.3 (“Bunny-Wunnies Freak Out”), via RStudio (Version 1.3.1093). Data were imported from excel using the Readxl package [20]. Where needed, data were selected with functions from the Dplyr package [21].

### Qualitative Comparative Analysis

We selected csQCA as it allows for more straightforward definitions and a conservative approach to calibration [18]. (In the field of data analysis, “conservative” means cautious to prevent false positive conclusions.) We performed two separate QCAs; the first for translational success, and the second for translational failure. All cases with calibrated translational success or failure (see below) were included in both QCAs.

### Data Calibration & data matrix

We calibrated all data to create so-called crisp sets as described in table 1. Data were calibrated on theoretical grounds and based on expert opinions from within our network. For example, cut-offs for old publication age at around the start of the century were based on the use of the internet becoming increasingly common in research. Also, >75 observations is considered a large study in the qualitative field. Data distributions were known at the time of calibration, and informed set definitions to the extent that (near) empty sets were consciously prevented.

**Table 1:** Data calibration for QCA

| Set name | Definition IN set | Definition OUT set |
| --- | --- | --- |
| Old | Publication date <2000 | Publication date ≥ 2000 |
| Intervention | "Interventions" were the unit of analysis | "events" or "studies" were the unit of analysis |
| Large | k>75 | K≤75 |
| MultSpec | At least two animal species were analysed | At most one animal species was analysed |
| Quantitative | Translation was calculated in a continuous manner | Translation was defined in a binary manner |
| Translation (outcome) | Success: >80% | Failure: <45% |

Units of analyses in the included reviews could be interventions, publications & studies, and particular (e.g., adverse) events. We distinguished observations at the intervention level from those at the event and study level, as the latter two are both chance processes, while at the intervention level we can imagine a clear distinction between e.g., compounds that do translate well and those that do not.

Translational success is difficult to define quantitatively [6]. For our QCAs, we selected the less disputable percentages only: success was defined as >80% correspondence, failure as <45%. We excluded the reviews with percentages from 45 to 80% (which were included in the RGA described below).

Set memberships scores were added to the data file in separate columns.

### Truth table creation & logical minimisation

A truth table was created with the truthTable function from the QCA package [22]. We analysed the truth table for sufficiency and necessity of individual factors before addressing combinations of factors. Sufficiency is the presence of the outcome in all cases with the occurrence of a predicting factor, and the factor is never present without the outcome (F → O, If F then O); necessity is the presence of a factor in all cases with the occurrence of an outcome, but the factor can also be present in cases without the outcome (F ← O, If O then F) Next, logical minimisation of configurations was performed with the minimize function from the QCA package.

We anticipated both logical inconsistencies and logical remainders in the truth table. Logical inconsistencies are rows with inconsistent outcomes (i.e., configurations that had both translational success and translational failure). This reanalysis of available data does probably not include all predictive factors relevant to translational success, which would result in perfectly defined sets without logical inconsistencies. Logical remainders are theoretically possible configurations that are not present in the data. Configurations with inconsistent translation and logical remainders were accepted, but they were not used to inform logical minimisation (in the QCA package’s truthTable function: incl.cut = 1, n.cut = 1, pri.cut = 0). Also, because of our awareness of missing relevant factors in this proof of concept study, we did not analyse coverage of the solutions; i.e., which part of the cases could be explained with the final formula.

### Regression analysis

All cases with complete data were included for the RGA. The following variables were included in the RGA: definition type (binary vs. continuous), unit of analysis (event, intervention or study), multiple species, the number of included observations and the year of publication. Compared to the QCA, we included more data into the RGA. Because we did not have to dichotomise data into crisp sets, we included all cases with full data, also those with translational success from 45% to 80%, with the original percentage as the outcome. The variables for the number of included observations and the year of publication were also included as numbers instead of dichotomising them. The variables definition type, unit of analysis and multiple species were included as binary variables, exactly like in the QCA.

Regression analysis was performed with the lm function from R’s basic stats package [19]. We tested a single model, including all variables included in the QCA individually. To provide a comparison with the QCA outcome we further added an interaction term for the variables “MultSpec” and “Quantitative”.

## RESULTS

### QCA

In our original review, we included 232 cases from 121 references. From these original cases, the 104 without missing data but with clearly successful or clearly unsuccessful translation were included in the QCA. Of these, 50 showed successful translation and 54 did not. The number of cases included in each set as defined in table 1 is shown in table 2.

**Table 2:** Number of cases per set

| Set name | N IN set (%) |
| --- | --- |
| Old | 30 (29%) |
| Intervention | 93 (89%) |
| Large | 15 (14%) |
| MultSpec | 26 (25%) |
| Quantitative | 73 (70%) |

The different observed set configurations with the outcomes are summarised in a truth table (table 3). The truth table shows that 9 configurations had inconsistent (both successful and unsuccessful translation) results. For 16 configurations, no cases were observed; these are the so-called logical remainders. None of the configurations was deemed implausible.

**Table 3:** truth table. Green: configuration consistent with translational success. Yellow: Configuration with inconsistent results. E.g., in configuration 2, the second line, the first one in yellow, there are 5 cases with this configuration of potential predictors, of which 4 show translational success, and 1 translational failure. Reddish: configuration consistent with translational failure. White: logical remainders.

| Configuration | Old | Intervention | Large | MultSpec | Quantitative | N cases (n successful translation) | Cases |
| --- | --- | --- | --- | --- | --- | --- | --- |
| 1 | 0 | 0 | 0 | 0 | 0 | 0 (0) | - |
| 2 | 0 | 0 | 0 | 0 | 1 | 5 (4) | Sultan_2017 (5x) |
| 3 | 0 | 0 | 0 | 1 | 0 | 0 (0) | - |
| 4 | 0 | 0 | 0 | 1 | 1 | 2 (2) | Yen_2014 (2x) |
| 5 | 0 | 0 | 1 | 0 | 0 | 0 (0) | - |
| 6 | 0 | 0 | 1 | 0 | 1 | 0 (0) | - |
| 7 | 0 | 0 | 1 | 1 | 0 | 2 (0) | Olson_2000 (2x) |
| 8 | 0 | 0 | 1 | 1 | 1 | 0 (0) | - |
| 9 | 0 | 1 | 0 | 0 | 0 | 10 (3) | Dong_2011, DeBuck_2007, Evans_2006, Sanoh_2014, Tang_2005, Wang_2010, Ward_2005, Ward_2008 (3x) |
| 10 | 0 | 1 | 0 | 0 | 1 | 33 (17) | Akabane_2010, Cao_2006, Cheng_2008 (2x), Chiou_2000a, Chiou_2000b (2x), Chiou_2002 (2x), Jones_2012 (2x), Grime_2013, Jones_2016 (4x), Kalvass_2007 (2x), Lennernas_2007, Ling_2009, Musther_2014, Paine_2011, Rocchetti_2007, Sanoh_2012, Walton_2004, Ward_2009 (2x), Whiteside_2008, Whiteside_2010, Wong_2004 (4x) |
| 11 | 0 | 1 | 0 | 1 | 0 | 4 (2) | Corpet_2005, Fagerholm_2007a, Fourches_2010, Goteti_2010 |
| 12 | 0 | 1 | 0 | 1 | 1 | 5 (4) | Mahmood_2001, Mahmood_2004, Mahmood_2013 (2x) Wajima_2002 |
| 13 | 0 | 1 | 1 | 0 | 0 | 7 (2) | Monticello_2017 (3x), Nagilla_2004, Weaver_2003 (3x) |
| 14 | 0 | 1 | 1 | 0 | 1 | 2 (0) | Musther_2014 (2x) |
| 15 | 0 | 1 | 1 | 1 | 0 | 4 (2) | Ennever_2003, Fourches_2010, Monticello_2017 (2x) |
| 16 | 0 | 1 | 1 | 1 | 1 | 0 (0) | - |
| 17 | 1 | 0 | 0 | 0 | 0 | 1 (0) | Litchfield_1961 |
| 18 | 1 | 0 | 0 | 0 | 1 | 0 (0) | - |
| 19 | 1 | 0 | 0 | 1 | 0 | 1 (0) | Steinberg_1987 |
| 20 | 1 | 0 | 0 | 1 | 1 | 0 (0) | - |
| 21 | 1 | 0 | 1 | 0 | 0 | 0 (0) | - |
| 22 | 1 | 0 | 1 | 0 | 1 | 0 (0) | - |
| 23 | 1 | 0 | 1 | 1 | 0 | 0 (0) | - |
| 24 | 1 | 0 | 1 | 1 | 1 | 0 (0) | - |
| 25 | 1 | 1 | 0 | 0 | 0 | 1 (0) | Schein_1973b |
| 26 | 1 | 1 | 0 | 0 | 1 | 19 (10) | Boxenbaum_1982 (4x), Chiou_1998 (2x), Crouch_1979 (2x), Fagerholm_1996, Freireich_1966 (4x), He_1998 (2x), Schneider_1999 (2x), Sietsema_1989 (2x) |
| 27 | 1 | 1 | 0 | 1 | 0 | 1 (0) | Schein_1973a |
| 28 | 1 | 1 | 0 | 1 | 1 | 7 (4) | Bachmann_1989, Mahmood_1996b, Mahmood_1996c, Mahmood_1998a, Mahmood_1998b (2x), Sietsema_1989 |
| 29 | 1 | 1 | 1 | 0 | 0 | 0 (0) | - |
| 30 | 1 | 1 | 1 | 0 | 1 | 0 (0) | - |
| 31 | 1 | 1 | 1 | 1 | 0 | 0 (0) | - |
| 32 | 1 | 1 | 1 | 1 | 1 | 0 (0) | - |

The configurations with inconsistent results indicates that none of the analysed factors was individually sufficient; successful (or unsuccessful) translation was not consistently present with occurrence of any of the analysed predictive factors. We further checked configurations for individual necessary conditions. None of the included factors was always present (or absent) when successful translation occurred, indicating that none of the included factors was individually necessary for translational success. Similarly, none of the included factors was always present (or absent) when translational failure occurred, so none of the individual factors was necessary for translational failure either.

Following our conservative approach excluding inconsistent configurations, the only consistent configuration corresponds with the solution from the logical minimisation process, i.e., the following formula for translational success:

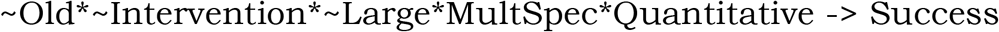

This means that the combination of relative recency (∼ means not), analyses at event or study level, n<75, inclusion of more than one species and quantitative analyses resulted in successful translation (>85%).

Further evaluation of the two cases consistent with this formula shows that they were both derived from the same publication; they are two meta-analyses (for different outcomes) including both animal and human data (further described in the discussion). The results from both meta-analyses showed a high degree of overlap between the animal and the human data.

A separate QCA on the reverse outcome results in the following formula for translational failure:

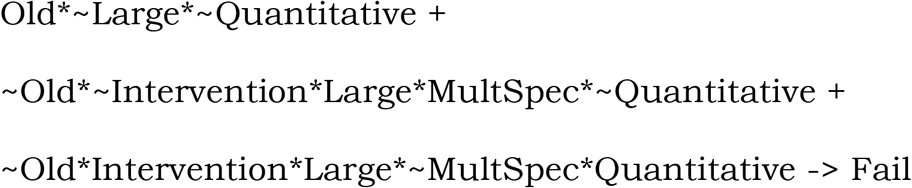

This formula arose from logical minimisation of the 6 configurations with a consistent negative outcome in table 3. It shows that there are 3 combinations of factors that consistently combine with translational failure; first, old, small studies using binary definitions of translation, second, newer large studies at the event or study level analysing multiple species using binary definitions of translation, and third, newer large studies at the intervention level analysing single species using quantitative definitions of translation.

### RGA

From the 232 cases from our original review, the 197 cases without missing data for the analysed variables were included in the RGA. The RGA included five explanatory variables, corresponding to the sets included in the QCA, and an interaction term. Two of these variables were numerical and three categorical-binary. The observed values of the variables are summarised in table 4.

**Table 4:**
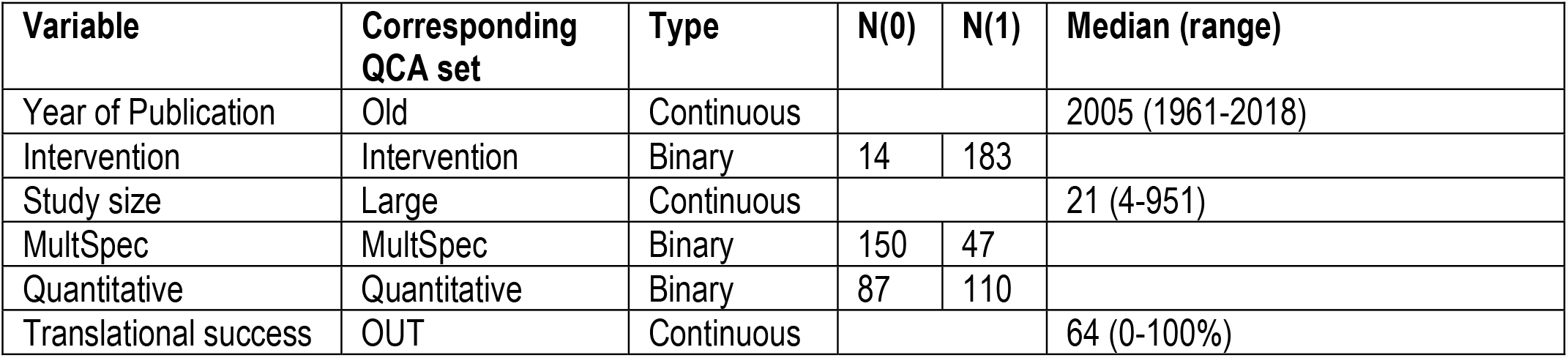
Summary of data included in the RGA

A summary of the RGA is provided in table 5. The effect estimates of the individual variables are relatively modest, but note that the effect sizes for publication age and study size are per year/ observation. The interaction term had a relatively large effect estimate, consistent with our QCA, indicating that analysing multiple species in combination with analysing translational success quantitatively may be optimal. However, none of the individual variables, nor this interaction, statistically affected translational success in the RGA.

**Table 5:** Summary of the RGA

| Variable | Estimate | SE | p |
| --- | --- | --- | --- |
| (Intercept) | -77.0 | 301.6 | 0.80 |
| Year of Publication | 0.07 | 0.15 | 0.65 |
| Intervention | -1.3 | 7.3 | 0.86 |
| Study size | -0.02 | 0.02 | 0.65 |
| MultSpec | 1.1 | 6.6 | 0.87 |
| Quantitative | 2.5 | 4.3 | 0.56 |
| MultSpec* Quantitative | 10.3 | 8.9 | 0.25 |

## DISCUSSION

We initiated these analyses as a first exploration and proof of principle of the QCA-method in meta-research of animal-to-human translation. QCAs have successfully been performed on data from systematic literature reviews in other fields [16, 17, 23, 24]. However, to the best of our knowledge, we are the first to perform a QCA with animal metadata, and to use it to analyse animal-to-human translation.

Our QCA resulted in a preliminary success formula for translational success at the meta-level; recent small reviews with analyses at the event or study level including more than one species and using a quantitative definition of translation were consistent with successful translation. While the effect sizes and directions of the RGA were consistent with these results, hence supportive of the QCA, the RGA did not identify any of the variables, nor the interaction term, as statistically significant. This shows the strength of the QCA approach.

### Cases consistent with the QCA-derived formulae

The formula for translational success was based on 2 meta-analyses, which both came from the same paper [25]. The authors performed an in-depth systematic review on guided tissue regeneration for periodontal infrabony lesions. They included 13 human and 9 animal papers, with varying study quality scores. The approach in their paper can be considered exemplary in synthesising animal and human data; combining them into a sub-grouped meta-analysis of percentages of bone filling, allowing for cross-species comparisons.

The formula for translational failure was based on 4 cases from 2 papers including newer studies [26, 27], combined with 4 papers combining into a single term for older studies [28-31]. To start with the newer studies; Olson et al. [27] described large analyses of animal studies in dogs, primates, rats, mice and guinea pigs. The authors were fairly optimistic in describing that 71% of human adverse events was somehow predicted in an animal model, but they also detailed low concordance rates in toxicity. Musther et al. [26] described correlational analyses of oral bioavailability, and concluded that bioavailability in animals is not predictive of that in humans. They provided separate data for mice (30 compounds), rats (122 compounds) and dogs (125 compounds), which were individually included in our analyses. Their monkey data (41 compounds) were included in the RGA, but excluded from the QCA because of an intermediate translational success rate.

To continue with the older studies; Litchfield [28] concluded that many serious side effects that can occur when a drug is given to humans were not predictable from observations on dogs or rats. The rat data were included in the QCA as clear translational failure, the dog data were only included in the RGA because of intermediate translational success. Steinberg & Schlesselman [31] compared the effects of pancreatitis therapeutics between 13 human studies and 25 animal studies in dogs, pigs, rats and guinea pigs, with low correspondence between the results.

Schein co-authored two publications in 1973 that both described multiple analyses included in our RGA, most with translational success rates between 45 and 80%. In one publication Schein and Anderson carefully concluded that combining data from multiple species could reduce false negatives for prediction of human adverse events, but one of their data sets reflected translation below 45% and was included in our QCA [29]. In the other publication, Schein et al. concluded that animal models can predict a substantial part of the adverse events occurring in clinical use [30], but again, translation was low in one of their data sets which we included in the QCA.

The term in the formula for translational failure that combines the 4 configurations listing these older studies (Old*∼Large*∼Quantitative) effectively illustrates the concept of logical minimisation, and thereby the potential of the QCA-method.

The formula we here present for translational success is restricted to smaller studies, and two of the three terms in our formula for translational failure cover large studies. While translational success being related to smaller studies may seem counterintuitive, we would like to mention that the familiarity with the data and the individual cases may be better with smaller studies, which might benefit the quality of the work, which, in specific configurations, could positively affect translational success.

### Suggestions for future QCAs

The here-presented data were not collected with QCA in mind, and therefore not optimised for this approach. However, they still resulted in preliminary formulae consistent with translational success and failure, based on relatively few consistent configurations. We expected contradictory lines in the truth table; configurations that were not consistent with the outcome. We hypothesise that this is mainly due to not all relevant factors being included in this QCA.

Future studies should gather data for more factors, but also on a larger number of cases to fill the logical remainders. A 2-step approach is considered; a first large QCA could comprise multiple factors relating to the meta-level. A second QCA could be restricted to the successful configurations from the first, and address factors at the primary study level. With more cases included, and less concern about logical remainders, multivalue QCAs (mv-QCAs) [32] may well be preferable. Concerning the currently included factors, it would be relevant to distinguish studies at the event and the study level instead of grouping them outside the set of studies at the intervention level. While it may seem like an attractive idea to include a factor for individual species, the resulting truth table would become incredibly large and have many logical remainders for the less frequently used species. However, a category distinguishing e.g., rodents, non-human primates and other mammals could be viable for future work.

QCAs can also be applied to other types of data than literature [14, 15, 18], which may make other types of data and variables accessible for analyses. E.g., individual compounds or targets can be defined as cases in a QCA, or communication and consideration of all available data in experimental design can be added as factors. There are indications that animal data are insufficiently considered in the design of human trials [33, 34], which might partially explain translational failure. Data could be gathered from multiple sources comprising also investigators’ brochures, ethics applications and patent registrations.

### Implications and conclusion

While our results are not conclusive and need confirmation, analysing multiple species in combination with analysing translational success quantitatively may be the optimal approach for future studies aiming to translate to humans. Analysing animal-to-human translation quantitatively as a percentage of correspondence instead of making simplified binary yes/no distinctions fairly reflects the available data. While we do not encourage to increase the number of animal studies overall, if a study aiming at translation is considered to be necessary, we may need to get used to the idea of testing more than one species.

In this paper, we present the first QCAs addressing translational success and failure rates. While the data were not collected with this method in mind, we show that the approach is viable, relevant and promising. Further knowledge on animal-to-human translation may help to improve efficiency in research and drug development, and to focus animal studies to where they are predictive.

## Data Availability

All data synthesised in the manuscript are available in the public domain

## Notes

### Competing Interest Statement

The authors have declared no competing interest.

### Funding Statement

ZonMW (40-42600-98-417), NWO (313-99-310) and the Federal State of Lower Saxony (R2N).

